# Classification of submandibular salivary stones based on ultrastructural studies

**DOI:** 10.1101/2020.10.20.20215822

**Authors:** Dmitry Tretiakow, Andrzej Skorek, Joanna Wysocka, Kazimierz Darowicki, Jacek Ryl

## Abstract

**Introduction:** Sialolithiasis remains a clinical problem with unclear etiopathogenesis, lack of prevention methods and only surgical treatment.

**Materials and Methods:** An ultrastructure examination of submandibular sialoliths obtained from patients with chronic sialolithiasis was conducted using a scanning electron microscope and X-ray photoelectron spectroscopy.

**Results:** Based on the results, we divided sialoliths into three types: calcified (CAL), organic/lipid (LIP) and mixed (MIX). The core structure of the CAL and MIX is very similar. The core of the LIP has a prevalence of organic components. The intermediate layers’ structure of the CAL is different from LIP and MIX. In LIP and MIX, the organic component begins to increase in intermediate layers rapidly. The structure of the superficial layers for all types of sialoliths is similar.

**Conclusions:** We introduced a new classification of the submandibular salivary gland stones. Based on the results, it can be said that sialoliths type CAL and LIP have their separate path of origin and development, while MIX is formed as CAL stone, and the further pathway of their growth passes as LIP stones. Organic components was much more than inorganic in all layers of salivary gland stones, which highly prevents their dissolution in the patient’s salivary gland duct.

## INTRODUCTION

Despite a large number of hypotheses, the mechanisms of formation and development of the sialoliths within the salivary gland ducts or/and parenchyma remain unexplained [1–5]. Lack of sufficient data on the etiopathogenesis of sialolithiasis does not allow causal treatment nor prevention. In most cases, surgical therapy is used [6–9].

We found that Raman spectroscopy allows for preliminary analysis of the structure of sialoliths with a qualitative but not quantitative assessment of their composition, which significantly reduces the research value of this method [10]. We decided to continue our research using spectroscopic analytical techniques, which offer not only the composition but also the chemical status of sialoliths. Such methods may include X-ray photoelectron spectroscopy (XPS).

Scanning electron microscopy (SEM) in the back-scattered electron (BSE) mode allows imaging and explicit differentiation of the salivary stone phases. Based on SEM, we noticed that sialoliths could also be analysed in terms of inorganic and mixed type, which may be useful in further research contribute to the classification of sialoliths in the future.

To broaden the knowledge about the etiopathogenesis of the sialolithiasis, we performed ultrastructural studies of separate layers of the salivary gland stones. Among numerous different types of chemical compounds recognized with high-resolution XPS, we have chosen some to act as markers for further classification. Several basic types of sialoliths can be distinguished despite the large variety of its structure.

## MATERIAL AND METHODS

We analyzed samples of 50 salivary sialoliths obtained from the submandibular gland main duct from 50 patients with submandibular sialolithiasis. The exemplary micrographs made using a SEM, operating with a variable pressure chamber at a pressure of 90 Pa to minimize ionization of the specimen. A back-scatter electron BSE detector was used for the analysis. The accelerating voltage was 20 kV. The only pre-treatment procedure was cutting the investigated salivary stones to reveal their core. Energy Dispersive X-Ray Spectroscopy (EDX) carried out using an UltraDry detector (ThermoFisher Scientific, United States) integrated with the SEM microscope.

The molecular composition of the specimens (X-Ray Photoelectron Spectroscopy, XPS) was studied using an Escalab 250Xi spectroscope (ThermoFisher Scientific, United Kingdom). The spectroscope was equipped with an Al Kα monochromatic X-Ray source, spot diameter 250 µm. The pass energy applied was 20 eV. Before measurements on sialoliths, the XPS was calibrated using single-crystal Au. Charge compensation was controlled through low-energy Ar+ ions emission utilizing a flood gun. The deconvolution procedure was performed using Avantage software (ThermoFischer Scientific).

The XPS analysis was carried out on 50 different salivary stones. For more detailed classification, each salivary stone was cut into pieces, taking a specimen from three different layers for further analyses: core, intermediate and the surface layer. This procedure is due to heterogeneity of sialoliths volume, as described in literature [10–12]. Thus, the averaged composition of ach different layer of each stone could be assessed for the very first time. High-resolution XPS spectra were recorded in a binding energy range of C 1s, O 1s, N 1s, P 2p, S 2p, Ca 2p, Na 1s, Mg 1s, Cl 2p, Si 2p and Fe 2p photopeaks. Up to three analyses were made for each specimen taken from each salivary stone layer, resulting in a dataset of over 200 independent specimen analyses.

The Shapiro-Wilk test confirmed the preliminary assumption about the lack of Gaussian distribution of the analyzed variables; therefore, Shapiro-Wilk test was used for nonparametric analyses. The Mann-Whitney U test (Z) used to verify intergroup differences. Finally, the existence of relationships between variables was verified using Spearman’s rho correlation (r).

## RESULTS

In our earlier research on sialolith ultrastructure, we distinguished 2 types of submandibular salivary gland stones: layered and homogeneous [10]. As a result of the present study, we assumed the hypothesis that the structure of sialoliths is more diverse and introduced a working classification of sialoliths to facilitate the description of further studies. We divided sialoliths into 3 types: calcified (CAL) (n=13/50, 26%), organic/lipids (LIP) (n=11/50, 22%) and mixed (MIX) (n=26/50, 52%).

The calcified (CAL) stones have the highest hydroxyapatite contribution. Thus they were chosen arbitrarily as the ones with Ca2p peaks not smaller than 12 at.% in the outer surface layer. On the other hand, the total share of all C1s components was expected to be the highest in salivary stones with the dominant contribution of the organic phase. Thus, we chose 50 at.% as the arbitrary threshold for the organic/lipid (LIP) type of classification. We noticed that there was a wide spectrum of salivary stones with uncertain classification, a feature visible in particular when performing the chemical analysis in the depth profile. This type of sialoliths was labelled as mixed (MIX) stones. The majority of MIX stone was characterized by a layered, heterogeneous structure, regardless of shape or size. Among the 26 mixed stones, 19 had layered structure and growth kinetics defined above. It allowed us to draw a conclusion regarding the third type of stone. This type of stone was characterized by the highest diversity, as visualized in the SEM micrographs.

The rough areas for specimen extraction were shown on the example of one of the stones (Fig. 1a). This type of stone classified as MIX. In this case, the border between the core layer, the interlayer, and the surface layer is also easily distinguishable, which is not always the case (Fig. 1b,c). The micrograph was taken using a BSE detector, meaning that the darker area of the interlayer indicated the layer is built of elements of lower atomic mass, suggesting the lower degree of calcification and higher organic content. This layer, when developed, is also characterized by the most complex topography. In case of all the LIP and in the majority of MIX stones with the heterogeneous structure, the interlayer was not as pronounced as in the case of CAL stones and the specimens for XPS studies had to be extracted more intuitively.

**Figure 1.**
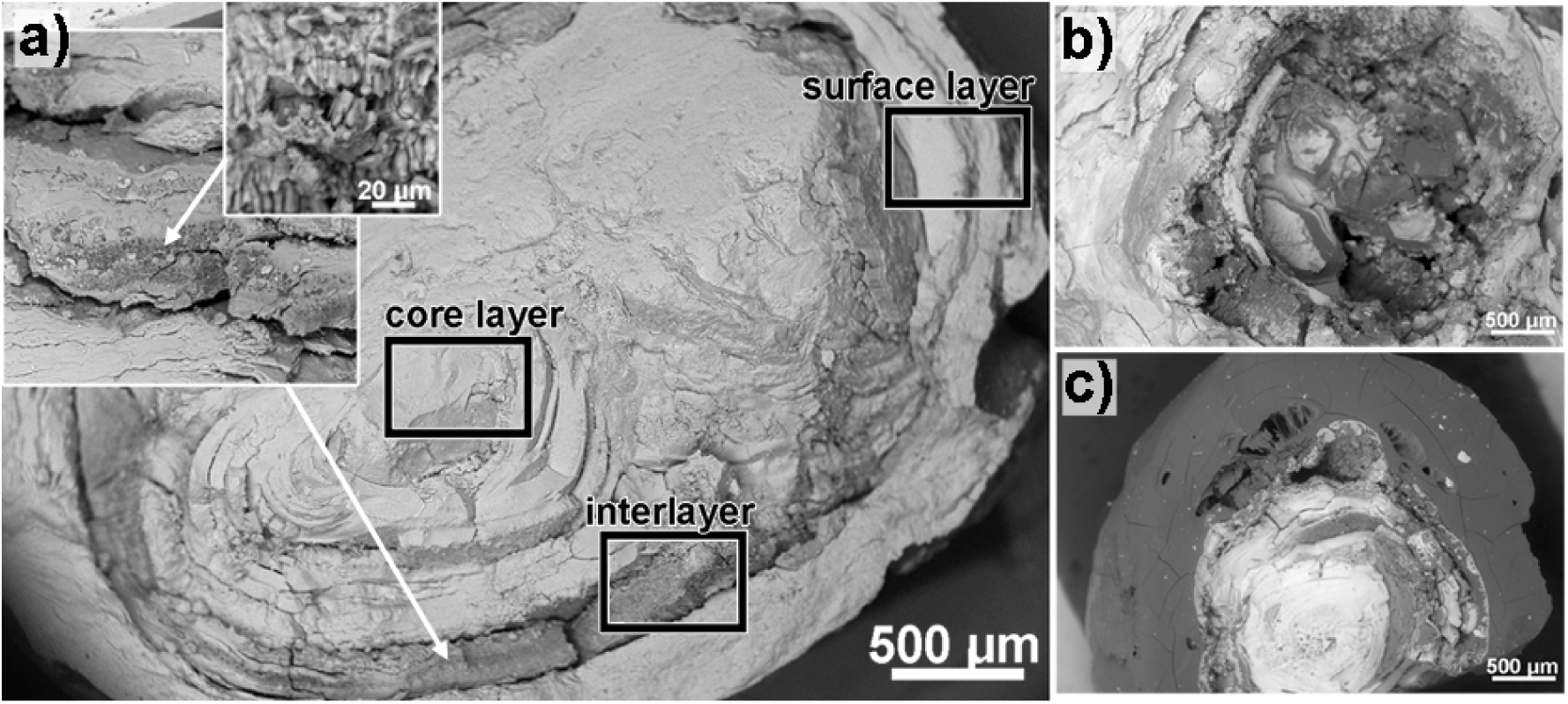
SEM backscatter images of three different salivary stones, classified as MIX type: a) reveals the areas of the core layer, interlayer, and surface layer for specimen extraction towards XPS analyses; b) and c) illustrate heterogeneity of chemical composition and structure between various MIX type stones (magnification x20; in the inset x120 and x750).

When carried out correctly, the high-resolution XPS analyses can provide deep insight into the surface chemical state of analyzed specimens. This action requires building a deconvolution model, indicating the peak binding energy (BE) for each identified chemical state. The spectra deconvolution based on the available literature survey and previously carried out our Raman spectroscopy studies [10]. Raman analyses confirmed the presence of various organic (collagen, glycoproteins, amino acids, carbohydrates) and inorganic compounds (hydroxyapatite, calcium phosphate, and magnesium phosphate, calcium carbonate).

Furthermore, once each chemical component were identified with the proposed deconvolution model, it was labeled as organic **{O}** or inorganic **{I}** material for further analyses. In the few cases where both organic and inorganic compounds were found within the studied BE, it was labelled as **{O/I}**. Figure 2 presents the exemplary and representative high-resolution XPS spectra recorded for the core layer, the intermediate layer, and the surface layer, revealing alteration in the chemical structures. These particular spectra recorded for one of the calcified stones. The spectral deconvolution was proposed, according to the model given below.

**Figure 2.**
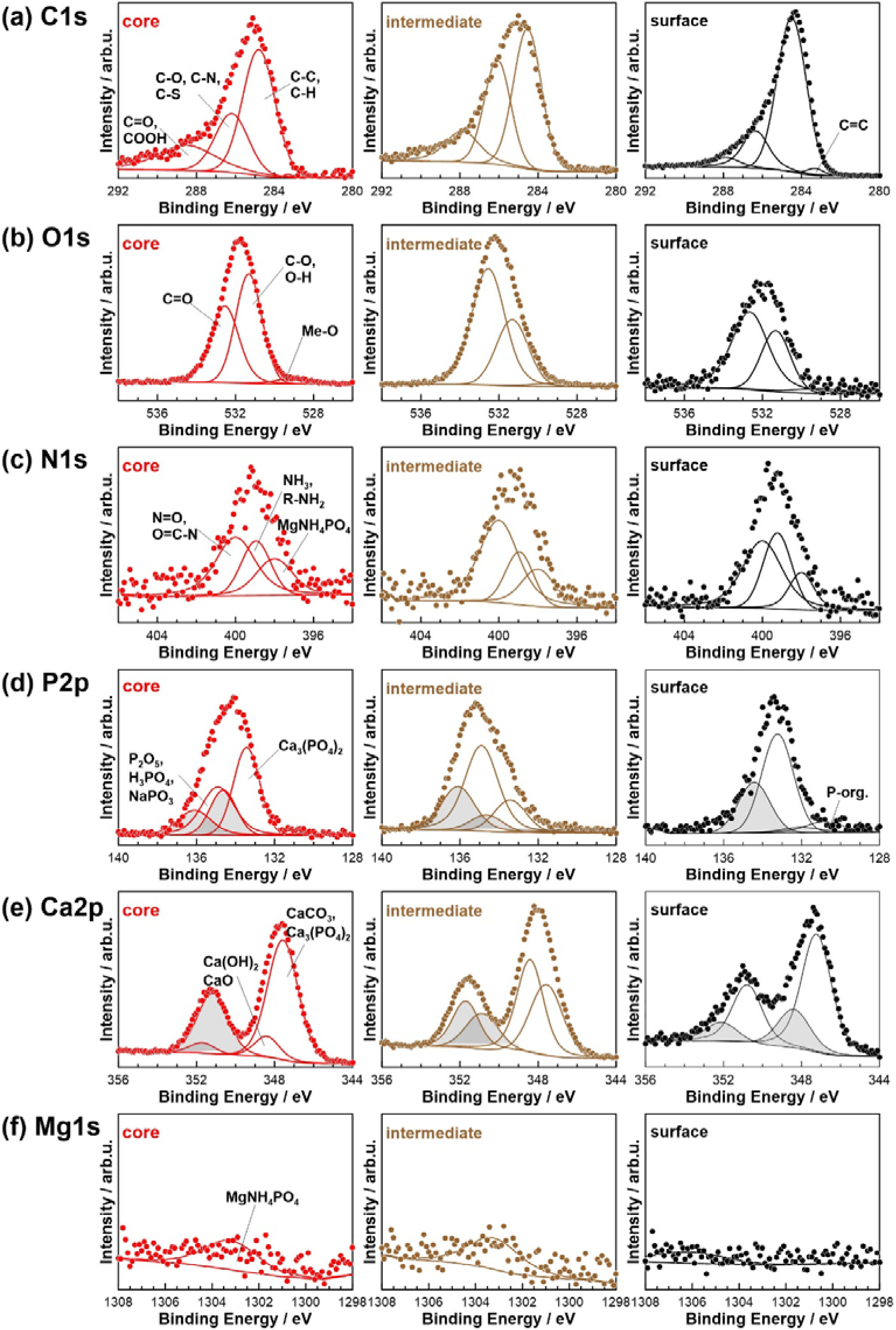
High-resolution XPS analysis was performed for a single representative calcified salivary stone, as an example of spectral deconvolution. The analysis was done in the (a) C 1s, (b) O 1s, (c) N 1s, (d) P 2p, (e) Ca 2p, and (f) Mg 1s photopeaks binding energy range. Specimens were taken from core, intermediate, and surface of calcified salivary stone layers.

Carbon has the largest share in the analyzed spectra, with at least four notable components to be found in the C 1s BE range. The primary peak, located at 284.6 eV, should be ascribed to the C-C and C-H bonds characteristic for hydrocarbon chains **{O}**. Two more peaks could be found at more positive energies, shifted by +1.6 and +3.5 eV vs. the primary component, respectively. Their position indicates the presence of C-O, C-N, C=N, and/or C-S bonds **{O}**. Carbonyl and carboxyl bonds within organic compounds typically manifest at the highest energy shifts, similar to inorganic carbonates, which often reported at approx. 288.1 eV. For this reason, the fourth peak classified as **{O/I}**. Finally, in some of the analyzed specimens, a small component was found at approx. 283.1 eV and characteristic for C=C bonds **{O}**. The analysis is in good agreement with the literature survey [13– 15].

The oxygen O 1s peak was also deconvoluted using four different components. Here, the organic/inorganic phase classification is more complicated and ambiguous. The most notable chemical component (532.7 eV) originates from C=O bonds that can be found in various organic compounds as well as inorganic carbonates **{O/I}**. The second strongly developed component, shifted by −1.5 eV, can be ascribed to organic C-O bonds but also various hydroxides **{O/I}**. Two smaller O 1s components were located at 529.4 and 534.9 eV. The first of the two should be assigned to metal-oxygen interaction **{I}** while the latter with N=O and/or carboxyl bonds **{O}**. A similar interpretation can be found in the literature [15,16].

Next, a phosphorous contribution observed in the form of three to four peak doublets. Unlike the previously examined elements, the major chemical state of phosphorous, with P 2p_3/2_ located at ∼133.3 eV, should be ascribed to an inorganic compound, namely Ca_3_(PO_4_)_2_ **{I}**. The second peak doublet with P 2p_3/2_ at 135.1 eV is characteristic for phosphorous acid, its anhydride, or other inorganic salts **{I}** [13,17]. Then, there is also a small contribution found at 137.1 eV in the case of some of the investigated specimens. The component was ascribed to MgNH_4_PO_4_*H_2_O **{I}**. The only plausible organic form of phosphorous was reported at approx. 130.6 eV and did not exceed 1 at.% in any studied layer, regardless of salivary stone classification.

Similar to phosphorous, calcium was found within salivary stones primarily in the form of inorganic compounds, deconvoluted within two peak doublets. The first, with Ca 2p3/2 at 347.3 eV, is characteristic for Ca_3_(PO_4_)_2_ **{I}**, while the latter was shifted by +1.4 eV and was linked with calcium hydroxides and carbonates **{I}** [18,19].

In sum, phosphorous, carbon, oxygen, and calcium constitute at least 90 at.% of any studied sample composition. The fifth most common element was nitrogen, contributing up to 8 at.%, depending on the sample. The deconvolution of N 1s XPS spectra performed with the use of three spectral components. The two of them are characteristic for various organic forms of nitrogen, amine, and nitrile groups **{O}** (at 399.1 eV) as well as oxidized N forms (at 400.3 eV). Third, the least prominent component located in the energy range characteristic for imines (397.6 eV); however, it is most likely connected with inorganic compound MgNH_4_PO_4_*H_2_O **{I}** due to the position of other characteristic peaks [20].

Magnesium is the last element to be found nearly in every analyzed specimen, yet its share was not exceeding 1 at.%. The recorded Mg 1s spectra were deconvoluted using a singular component peaking at approximately 1303.1 eV. This peak is often ascribed as inorganic MgNH_4_PO_4_*H_2_O **{I}**. The remaining studied elements, e.g. sodium and chlorine, were found in small amounts, rarely exceeding 0.5 at.%, values too low for appropriate statistical analysis. Their peak position was characteristic for various inorganic salts. As such, these signals may originate both from salivary stones as well as result of artificial contamination. Similar may be said about sulphur. Its amount was found the highest in MIX and LIP sialolith. However, the sulphur share within analyzed spectra never exceeded 0.5 at.%. Interestingly, sulphur was found at BE of 163.4 eV, which is characteristic for organic compounds, e.g. thiols [21].

Apart from the above-presented observations, the high-resolution XPS data allows detailed statistical analysis. The in-depth quantitative analysis allowed to build simplified depth profiles, revealing chemical distribution within the volume of salivary stones. The total share of primary contributing elements was presented in Fig. 3 (r = 0.3; p <0.05).

**Figure 3.**
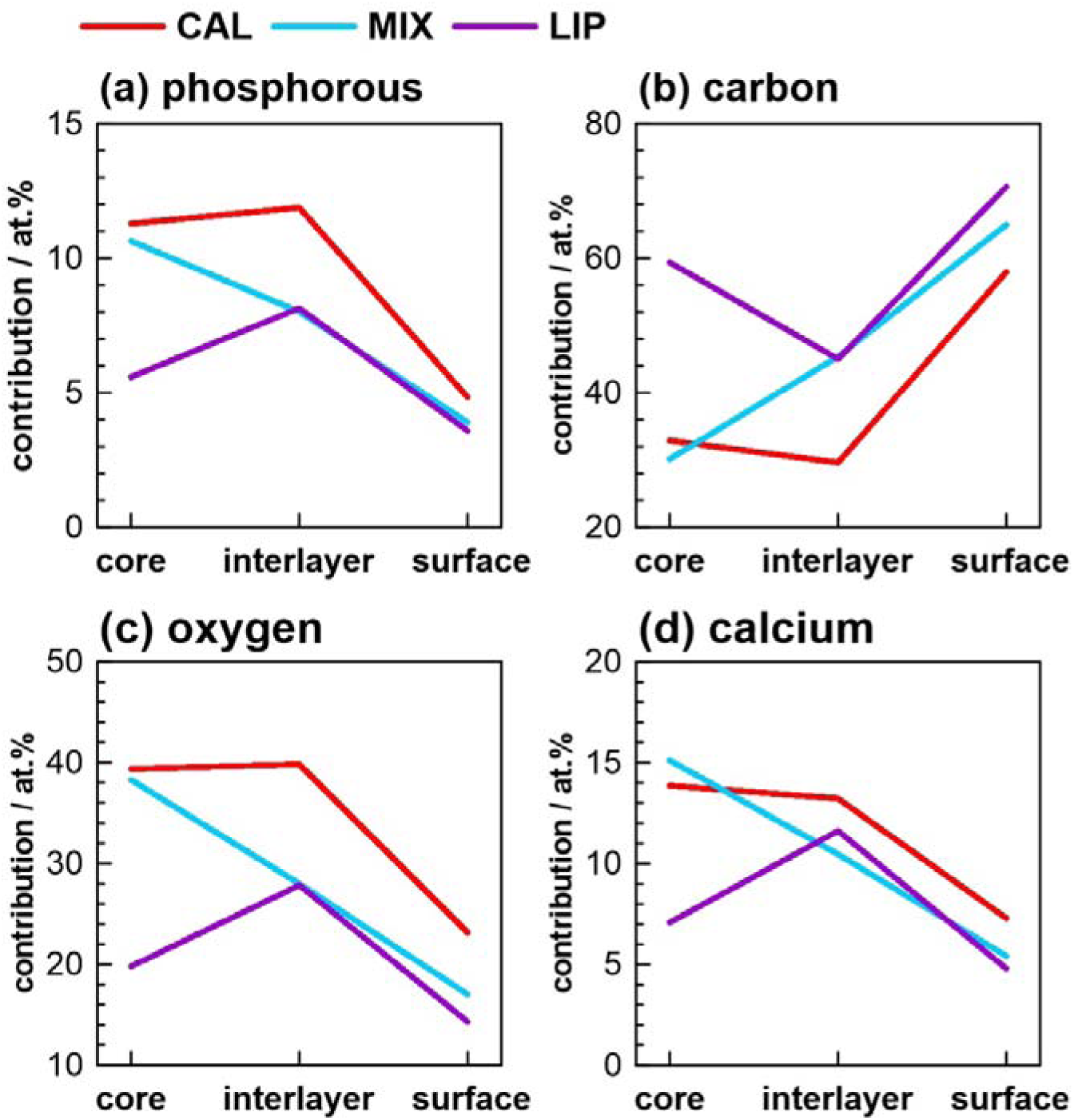
High-resolution XPS quantitative analysis of total contribution for the four primary components: (a) phosphorous, (b) carbon, (c) oxygen, and (d) calcium.

Next, the exemplary high-resolution XPS spectra, recorded in the binding energy of the core level P2p, Ca2p, and C1s photoelectrons, are demonstrated in Fig. 4a, b, c, respectively. This data reveals the differences in the chemistry of salivary gland stones based on their classification, but also the differences between the sialolith core and the surface, which is an extension of Fig. 1. The dashed lines indicate the peak position of various chemical compounds used for peak deconvolution according to the previously presented fitting model. It should be noted that there are small structural differences between the various analyzed sialoliths, even within the proposed classification type.

**Figure 4.**
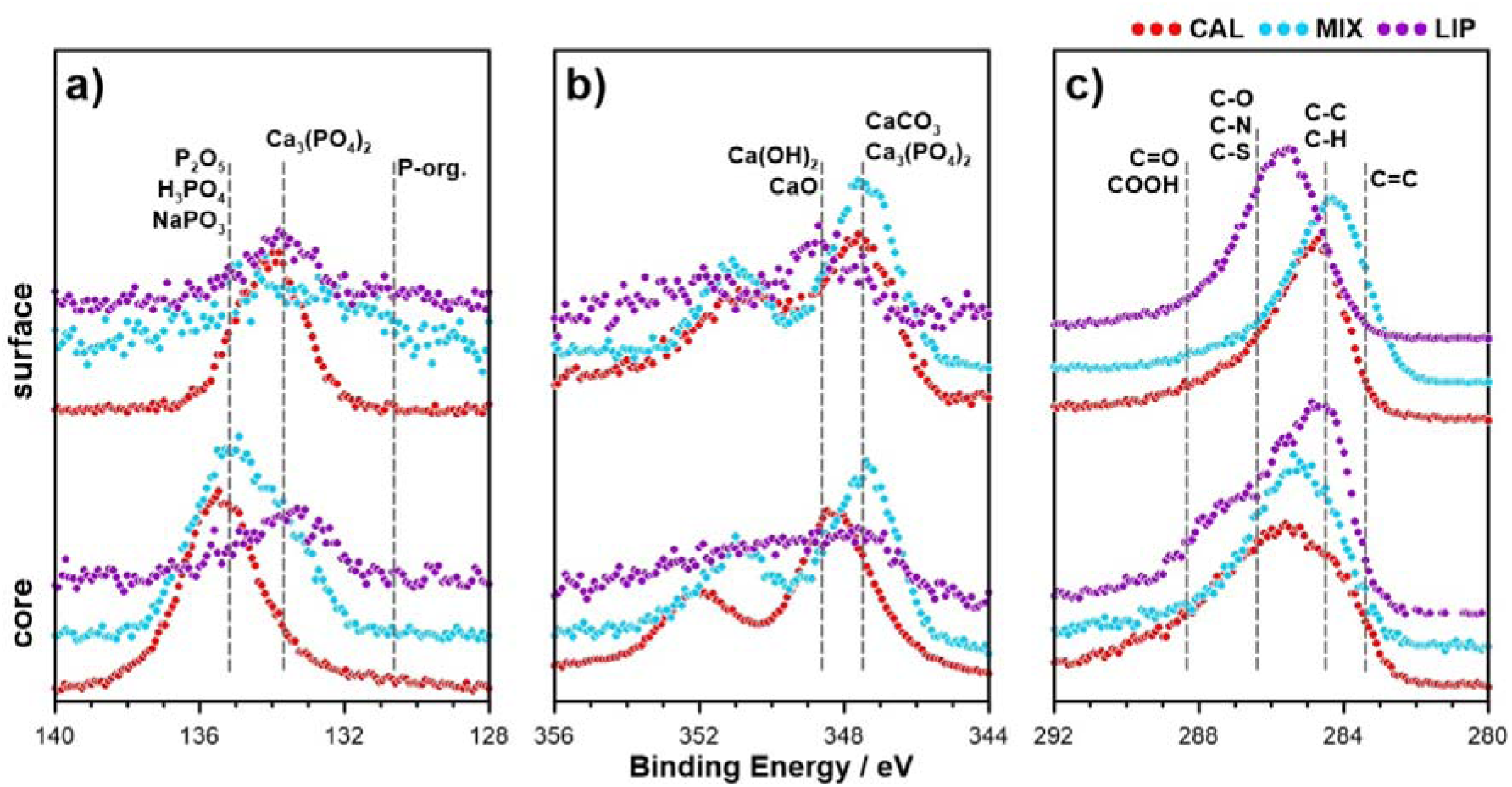
High-resolution XPS analysis of the salivary gland stones taken from the core and the surface layers of the specimens, depending on the proposed classification type. The analysis was carried out in the a) P2p, b) Ca2p, c) C1s binding energy range.

Fig. 5 represents the organic to inorganic phase contribution depending on the suggested classification type of sialolith and each analyzed layer. The analysis was performed based on previously-ascribed organic or inorganic labels {O} and {I}, with few disputable {O/I} cases diversified based on the assumed probability of the occurrence of given types of compounds. Analyzes have shown significantly differentiates the studied groups of sialoliths’ layers (Z = 107.0; p <0.05).

**Figure 5.**
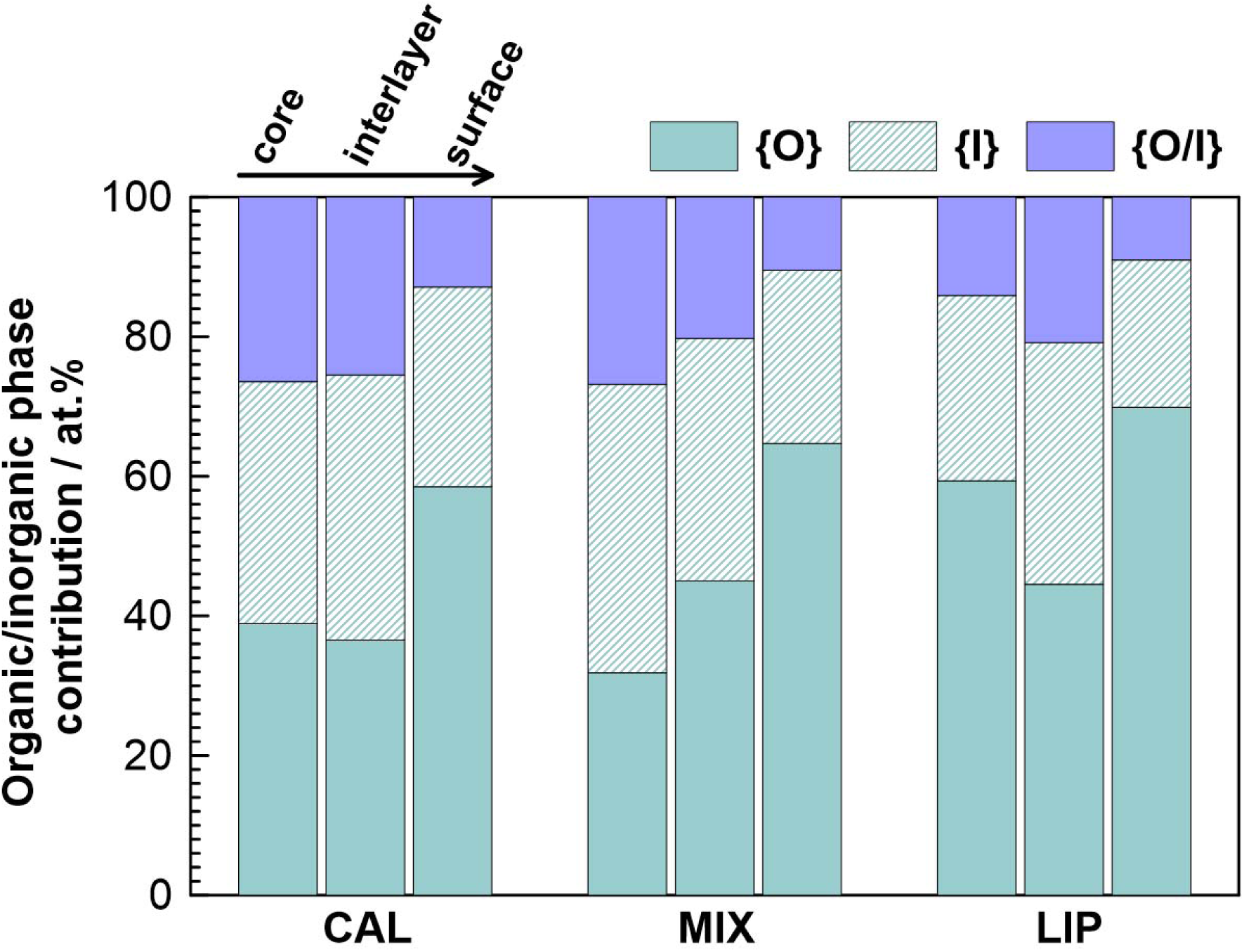
Organic to inorganic phase contribution (in at.%), based on high-resolution XPS studies of each type of sialolith and divided into three layers: core layer, interlayer, and surface layer.

## DISCUSSION

The cause of sialolithiasis has been researched for over 100 years. Unfortunately, there are still many aspects that remain unclear. The clinicopathologic investigations help in understanding of the etiopathogenesis of salivary gland stones.

In some studies the sialolith specimen had a core almost exclusively composed of organic material, and the core was surrounded by a highly mineralized shell structure [22]. In our study, in each group of sialoliths, the core consisted of an organic phase {O} and an inorganic phase {I} (Fig. 4). In LIP sialoliths type, the organic phase of the sialolith core reached up to 80%, which could be interpreted by some researchers as entirely organic [12,22]. Several studies describe sialoliths containing mainly inorganic components [2,23,24], which could correspond to CAL sialoliths in our study, where the inorganic phase consists of about 40%.

A few conclusions may be drawn based on the data analysis with the above-presented deconvolution model. As a general feature, the compounds containing aliphatic C-C bonds, esters and amines were notably the most dominant at the surface of any analyzed specimen, regardless of their proposed classification. However, the LIP sialoliths generally contain a higher amount of compounds with C-O, C-N, and C=O bonds. When comparing C1s spectra, it was observed that the highest amount of species with unsaturated bonds (C=C) was found in the interlayer of calcified specimens. It should be noted that in the case of CAL sialoliths, the highest amount of calcium phosphates was also found in the intermediate layers, where Ca 2p_3/2_ contribution reached as high as 8 at.%. Importantly, the highest amount of calcium can be found in the core of CAL sialoliths. Yet, its nearly equally high presence in stones with mixed classification may suggest that this type of stone originates from the CAL type of core. It should also be noted that calcium chemistry reveals certain differences, and the core of mixed-classified stones possesses a higher share of CaO and/or Ca(OH)_2_ in comparison with CAL. It is the fact that the level of the calcium in the submandibular salivary gland parenchyma is high. It is released when secretory granules disintegrate during secretory inactivity to precipitate on exposed phospholipid of damaged membranes or to stagnate in the lumina and precipitate there on exposed phospholipid [25].

The strength of the O 1s signal in either observed chemical state is typically better represented in the core layer and its share starts to decrease towards the surface layers. Furthermore, the lowest oxygen share was typically found within LIP specimens. Similarly, the amount of phosphates was decreasing in the surface layer, and its contribution was typically found the lowest in LIP classified specimens. It was also observed that the amount of primary and secondary amines was higher for LIP and MIX sialoliths rather than the CAL stones. Nevertheless, their amount does not significantly depend on the in-depth of analysis. Similar observations were described by other authors, where the core was mainly composed of carbon and oxygen, with less amount of nitrogen and a trace amount of sulphur [12,22,26].

Visualizing the quantitative comparison of each of the four primary elements in particular specimen layers leads to observable differences in the chemical analysis depending on the approached classification. In particular, it should be noted that within the inner core layer, the contribution of primary elements such as phosphorous, carbon, oxygen, and calcium of MIX sialoliths is nearly the same as in CAL ones (Fig. 3). Excluding carbon species, the concentration of these primary elements in LIP is significantly lower (p<0.01). Judging from the data presented in Fig. 4a and b, one can observe chemical similarities between the shapes of the core layers high-resolution P 2p and Ca 2p XPS spectra for CAL and MIX sialoliths.

In the outer surface layers, the chemistry of MIX sialoliths becomes significantly altered, and much more resembles the chemistry of LIP stones (see Fig. 3). A closer comparison of the spectral shape reveals some differences, though, such a higher share of calcium carbonates and phosphates in the MIX stones, and a higher share of carbonyls, esters, and amines in the LIP stones (see Fig. 4b, c). This observation indicates that both MIX and CAL stones may have a similar origin, but due to various environmental factors affect their growth and development have led to diversification.

In general, lipids constitute around 10% of the matrix weight of sialoliths, and most of them are neutral lipids, glycolipids, and phospholipids [22,27]. Lipid components are generally suspected to originate from the degenerating cellular membranes of the salivary gland cells [28]. There is also a significant differentiation to be made between various salivary stone types when referring to the share of their organic and inorganic components (Fig. 5). In general, the amount of organic phase is the highest at the surface of the analyzed specimens. However, it is visible that the chemistry in each sialolith type is different, and for the LIP stones, the segregation is less notable. These were also the smallest stones in general, where the interlayer was difficult to differentiate with the naked eye.

On the other hand, the above mentioned relationship is well developed for CAL and MIX types of stones. Furthermore, LIP stones possess much less inorganic phase, barely exceeding 20 at.%, while its amount is expectedly closer to 40 at.% for calcified stone specimens. It may indicate other causes of the stones formation and assume that may also result from bacteria or their biofilms. However, a proposal that immune cells interact with biofilm and initiate sialolith formation is not clear.

The intermediate layers’ structure of the CAL is different from LIP and MIX sialoliths. It may indicate the formation of sialoliths by depositing calcium microcrystals (sialomicroliths) on the previously formed core. In LIP and MIX sialoliths, the organic component begins to rapidly increase in intermediate layers, which may be the result of the development of local inflammation and intensification of a degenerating process of cellular membranes in the lumen of the main salivary gland duct.

A similar structure of the superficial layers of all types of stones may indicate a universal process of sialolith growth, in the phase of clinical manifestation (total obstruction of the salivary gland duct), but this is only our consideration.

The obstruction of the submandibular gland duct, alteration of secretory activity of the salivary gland or a change in the chemical composition of saliva (due to diet change, dehydration, accompanying diseases) lead to sialomicrolith deposition in salivary gland duct [26,29]A and probably cause of CAL core formation. The MIX and LIP sialoliths may arise due to infection or possibly bacterial biofilm within the submandibular gland duct and/or damage to the epithelial layer of the submandibular gland. The enlargement of these types of stones occurs in a slightly different way, which may be related to the severity of the inflammation or the body’s immune response.

According to Fig 5., all layers of salivary gland stones contain more organic components than inorganic, which may be the reason why they do not dissolve in the patient’s salivary gland duct. It is important to note that not all MIX stones have a clear classification, as was mentioned in our previous study (Tretiakow et al. 2020) and by other authors. On the other hand, our depth profile analysis revealed that the majority of sialolith classified as MIX have similar and specific growth kinetics, where their core is highly resembling the CAL stones. At the same time, while the surface layer is calcified, the chemistry of other primary components reveal observable differences. The content of inorganic compounds in salivary glands may predispose to the development of new non-invasive destruction methods of these deposits.

There is currently a lack of conservative treatment and methods of preventing sialolithiasis of the large salivary glands. Previous studies of sialoliths ultrastructure have shown their diversity, suggesting differences in the mechanisms of their creation and development. Creating a classification of sialoliths allows differentiating the scientists’ approach to modelling sialolithiasis on an animal model. It can be helpful in further studies of the etiology of salivary gland stones and understanding the factors that predispose them.

One of the limitations of the proposed approach is that our classification (LIM, CAL, MIX) might be too narrow. Specifically, the MIX-classified stones chemistry is varied, several sialoliths in the MIX group had a very unique composition and perhaps should be categorized separately. Another limitation is the XPS technique itself, which is highly surface-sensitive, thus the analysis results may be interfered by presence of the adventitious carbon due to air exposure of studied specimens. On the other hand, XPS provides deep chemical analysis, offering information about the chemical affinity, which is unique in comparison with any other surface analysis technique.

It would be beneficial if future studies focused on the origin and the chemical affinity of elements less commonly found in sialoliths e.g. sulphur or nitrogen. Based on the above discussed limitations, it would be highly valuable to extend current studies with more in-depth analysis of MIX sialoliths, make an attempt to broaden the sialolith classification.

## CONCLUSIONS

Based on the results, we introduced a new classification of the submandibular salivary gland stones: calcified (CAL), organic/lipids (LIP), and mixed (MIX).The structure of the CAL and MIX sialoliths core is very similar, which may suggest a similar mechanism for the origin of these types of sialoliths, unlike LIP, which has a different core structure. The structure of the intermediate layers is very similar in LIP and MIX type sialoliths, which may suggest a similarity of the mechanism of enlargement of these types of stones. The structure of the superficial layers for all types of sialoliths is similar, which may suggest a universal mechanism of stone enlargement at the stage of total impairment of the submandibular salivary gland duct. Based on the results obtained, it seems that sialoliths type CAL and LIP have a separate path of origin and development, while sialoliths type MIX formed as CAL stones, and the further path of their growth passes as LIP stones. However, this theory requires confirmation in further research on more material. Organic components were much more than inorganic components in all layers of sialoliths, which highly prevents their dissolution in the patient’s salivary gland duct.

## Data Availability

I declare the availability of all the data referred to in the manuscript. I provide the data on request.

## ACKNOWLEDGMENTS

This work was supported by departmental funding of the Faculty of Medicine (Medical University of Gdansk) and Faculty of Chemistry (Gdansk University of Technology). The study protocol was approved by the Regional Bioethics Committee of Gdansk Medical University, Poland (approval nr. NKBBN/452/2019). The authors declare no potential conflicts of interest concerning the authorship and/or publication of this article.

## Author contributions

Dmitry Tretiakow: contributed to study concept and design, contributed to acquisition, drafted manuscript, critically revised manuscript, gave final approval.

Andrzej Skorek: contributed to conception and design, contributed to acquisition, critically revised manuscript, gave final approval.

Joanna Wysocka: contributed to acquisition, analysis, and interpretation; drafted manuscript; gave final approval.

Kazimierz Darowicki: contributed to conception, critically revised manuscript, gave final approval.

Jacek Ryl: contributed to design, contributed to acquisition, analysis, and interpretation, drafted manuscript, critically revised manuscript, gave final approval.

All authors gave their final approval and agree to be accountable for all aspects of the work.

